# Early, low-dose and short-term application of corticosteroid treatment in patients with severe COVID-19 pneumonia: single-center experience from Wuhan, China

**DOI:** 10.1101/2020.03.06.20032342

**Authors:** Yin Wang, Weiwei Jiang, Qi He, Cheng Wang, Baoju Liu, Pan Zhou, Nianguo Dong, Qiaoxia Tong

**Affiliations:** Department of Cardiovascular Surgery, Union Hospital, Tongji Medical College, Huazhong University of Science and Technology, Wuhan, China; Department of Infectious Diseases, Union Hospital, Tongji Medical College, Huazhong University of Science and Technology, Wuhan, China; Department of Gastroenterology, Union Hospital, Tongji Medical College, Huazhong University of Science and Technology, Wuhan, China; Department of Rheumatology, Union Hospital, Tongji Medical College, Huazhong University of Science and Technology, Wuhan, China; Department of Hand Surgery, Union Hospital, Tongji Medical College, Huazhong University of Science and Technology, Wuhan, China

**Author notes:** Correspondence authors **Corresponding Author:** Nianguo Dong, M.D, Ph.D., Address: Jiefang Avenue 1277#, Wuhan, Hubei, 430000, China, Institution: Department of Cardiovascular Surgery, Union Hospital, Tongji Medical College, Huazhong University of Science and Technology, China, Qiaoxia Tong, M.D, Ph.D., Address: Jiefang Avenue 1277#, Wuhan, Hubei, 430000, China, Institution: Department of Infectious Diseases, Union Hospital, Tongji Medical College, Huazhong University of Science and Technology, China.

## Abstract

**Background:** Severe patients with 2019 novel coronavirus (2019-nCoV) pneumonia progressed rapidly to acute respiratory failure. We aimed to evaluate the definite efficacy and safety of corticosteroid in the treatment of severe COVID-19 pneumonia.

**Methods:** Forty-six hospitalized patients with severe COVID-19 pneumonia hospitalized at Wuhan Union Hospital from January 20 to February 25, 2020, were retrospectively reviewed. The patients were divided into two groups based on whether they received corticosteroid treatment. The clinical symptoms and chest computed tomography(CT) results were compared.

**Results:** A total of 26 patients received intravenous administration of methylprednisolone with a dosage of 1-2mg/kg/d for 5-7 days, while the remaining patients not. There was no significant difference in age, sex, comorbidities, clinical or laboratory parameters between the two groups on admission. The average number of days for body temperature back to the normal range was significantly shorter in patients with administration of methylprednisolone when compared to those without administration of methylprednisolone (2.06±0.28 vs. 5.29±0.70, P=0.010). The patients with administration of methylprednisolone had a faster improvement of SpO2, while patients without administration of methylprednisolone had a significantly longer interval of using supplemental oxygen therapy (8.2days[IQR 7.0-10.3] *vs*. 13.5days(IQR 10.3-16); P<0.001). In terms of chest CT, the absorption degree of the focus was significantly better in patients with administration of methylprednisolone.

**Conclusion:** Our data indicate that in patients with severe COVID-19 pneumonia, early, low-dose and short-term application of corticosteroid was associated with a faster improvement of clinical symptoms and absorption of lung focus.

## Introduction

Corona Virus Disease 2019 (COVID-19) was first reported in late December 2019, originating in Wuhan, China and then transmitted in Hubei Province, throughout China and even other countries[1,2]. This ongoing outbreak has already been declared by World Health Organization(WHO) as a Public Health Emergency of International Concern (PHEIC). As of February 28, there are 78,824 confirmed cases, 7,952 severe cases and 2,788 confirmed deaths across China[3]. COVID-19 is a beta-coronavirus, presenting as viral pneumonia with high infectiousness through respiratory droplets or direct contact and appears to have greater infectivity and a lower-case fatality rate when compared to severe acute respiratory syndrome (SARS) and Middle Eastern respiratory syndrome(MERS)[4,5]. Moreover, the pathological features of COVID-19 pneumonia greatly resemble those seen in SARS and MERS, and show diffuse alveolar damage with cellular fibromyxoid exudates, desquamation of pneumocytes and hyaline membrane formation, indicating acute respiratory distress syndrome(ARDS)[6]. Corticosteroid are widely used as therapy for ARDS and have been used to treat SARS with proof of efficacy, which could significantly decrease the mortality of severe SARS patients[7,8]. However, there is no evidence-based medical evidence to confirm the definite efficacy and safety of corticosteroid in the treatment of severe COVID-19 pneumonia. To address this issue, we have performed a retrospective study on the clinical and radiographic outcomes of treatment with or without corticosteroid for a cohort of patients with severe COVID-19 pneumonia.

### Patients and methods

We studied patients with severe COVID-19 pneumonia from January 20 to February 25, 2020 at the isolation ward of Union Hospital of Huazhong University of Science and Technology. Union Hospital is sentinel general hospital located in the endemic areas of COVID-19 in Wuhan, Hubei Province, and is designated by the government for the treatments of COVID-19 infection. The study was approved by the Research Ethics Committee of Tongji Medical College, Huazhong University of Science and Technology, Wuhan, China. Verbal consent was obtained from patients before the enrollment.

Diagnosis of 2019-nCoV pneumonia in this study were diagnosed according to the WHO interim guideline[9]. Throat swab sample of the patients was collected and COVID-19 was confirmed by real-time RT-PCR assay using a 2019-nCoV nucleic acid detection kit according to the manufacturer protocol (Shanghai bio-germ Medical Technology Co Ltd)[10]. The clinical classification according to the coronavirus pneumonia diagnosis and treatment plan (trial version 5) developed by the National Health Committee of the People’s Republic of China, and those meet any of the followings were diagnosed as severe case: (1) respiratory distress, respiratory rate per min≥30; (2) in the resting state, means oxygen saturation≤93%; (3) arterial blood oxygen partial pressure/oxygen concentration≤300mmHg (1mmHg = 0.133kPa). (4) other conditions such as older than 60 years, with complication of hypertension, diabetes, coronary disease, cancer, pulmonary heart disease, structural lung disease and immunosuppressed individuals.

The demographic characteristics, comorbidities, symptoms, clinical parameters, laboratory indexes and chest computed tomographic (CT) scan findings were extracted from electronic medical records. The vital signs of the patients (temperature, heart rate, blood pressure, respiratory frequency and oxygen saturation [SpO_2_]) were monitored daily; The blood tests (complete blood count, liver and renal function, C-reactive protein, Inflammatory factors and lactate dehydrogenase) were examined every 3 days; The chest CT scan was performed every 7-10 days. Assessment criteria of this study was the dynamic changes of the indexes mentioned above.

The Statistical Packages for Social Sciences, version 19.0 (SPSS) were used for data analysis with P < 0.05 as the criterion for significance. Continuous variables were reported as means and standard deviations, or medians and interquartile range (IQR) values. Categorical variables were reported as counts and percentages. Continuous variables were compared using Student’s t-tests, Mann-Whitney U test and categorical variables were compared using Fisher’s exact tests.

## Results

A total of 46 hospitalized patients with severe COVID-19 pneumonia were diagnosed and included in this study, the age, sex, comorbidities, clinical and laboratory parameters on admission for the are shown in Table 1. Their average age was 54 years (IQR 48-64), and 26 (57%) were males. Of these patients, 18 (32.1%) had at least one of the following underlying disorders: hypertension (14[30.8%]), diabetes (4[8.7%]), cardiovascular disease (6[13.0%]), Chronic pulmonary disease(3[6.5%]), cerebrovascular disease(2[4.3%]), and malignancy (2[4.3%]). The median and interquartile range of the main clinical and laboratory parameters are as following: body temperature 37°C(36.8,38.4) °C, heart rate 86bpm (74,96), BP 132(121,141)/77(70,85)mmHg, respiratory frequency 26bpm(21,31), SpO2 at rest 91% (86,92), and white blood cell(WBC) 7.74(5.24,9.96) G/L, neutrophil(PMN) 5.53(3.85,7.87) G/L, lymphocyte(LYM) 0.86(0.58,1.15) G/L, C-reactive protein(CPR) 61.5(14.4,109.9) mg/L, procalcitonin(PCT) 0.55(0.14,1.09) mg/L, interleukin-2(IL-2) 2.77(2.32,3.55) pg/ml, interleukin-4(IL-4) 2.17(1.87,2.61) pg/ml, interleukin-6(IL-6) 18.4(10.4,53.0) pg/ml, interleukin-10(IL-10) 6.7(3.9,8.1) pg/ml and ferroprotein(FER) 787(300,1057) µg/L. Three(5.4%) patients died during the hospitalization: a 89 years old man with the end stage of prostate cancer died because of multiple organ failure; a 51 years old woman with diabetic foot and gangrene died at 2 days after the amputation surgery; a 49 years old man died of respiratory failure. All of the other 43 patients were successfully cured and discharged.

**Table 1.** Clinical characters and main laboratory parameters of 46 patients with severe COVID-19 pneumonia

|  | All patients | Methylprednisolone treatment |  |  |
| --- | --- | --- | --- | --- |
|  | (n=46) | Yes(n=26) | No(n=20) | P value |
| Age, years | 54(48,64) | 54(48,63) | 53(48,63) | 0.916 |
| Male | 26(57) | 16(62) | 10(50) | 0.434 |
| **Comorbidity** |  |  |  |  |
| Chronic cardiac disease, n(%) | 6(13) | 3(12) | 3(15) | 0.729 |
| Chronic pulmonary disease, n(%) | 3(6.5) | 1(3.9) | 2(10) | 0.402 |
| Cerebrovascular disease, n(%) | 2(4.3) | 0(0) | 2(10) | 0.099 |
| Malignancy, n(%) | 2(4.3) | 1(3.9) | 1(5) | 0.849 |
| Diabetes | 4(8.7) | 3(12) | 1(5) | 0.435 |
| Hypertension, n(%) | 14(30) | 8(31) | 6(30) | 0.955 |
| **Clinical parameters on admission** |  |  |  |  |
| Temperature, °C | 37.6(36.8,38.4) | 37.6(36.7,38.0) | 38.2(36.8,38.6) | 0.221 |
| Heart rate, per min | 86(74,96) | 86(69,97) | 86(82,96) | 0.598 |
| SBP, mmHg | 132(121,141) | 135(121,140) | 128(120,144) | 0.584 |
| DBP, mmHg | 77(70,85) | 77(70,85) | 77(70,86) | 0.438 |
| Respiratory frequency, per min | 26(21,31) | 28(21,36) | 24(20,30) | 0.039 |
| SpO<sub>2</sub> at rest, % | 91(86,92) | 91(88,92) | 90(85,92) | 0.206 |
| **Laboratory parameters on admission** |  |  |  |  |
| WBC, G/L | 7.74(5.24,9.96) | 7.47(4.91,10.31) | 7.83(5.44,9.65) | 0.979 |
| PMN, G/L | 5.53(3.85,7.87) | 5.59(3.59,8.27) | 5.35(3.85,7.87) | 0.823 |
| LYM, G/L | 0.86(0.58,1.15) | 0.77(0.50,1.14) | 0.91(0.68,1.22) | 0.176 |
| CPR, mg/L | 61.5(14.4,109.9) | 78.9(33.7,115.6) | 61.3(15.7,109.9) | 0.646 |
| PCT, mg/L | 0.55(0.14,1.09) | 0.60(0.15,1.09) | 0.29(0.13,1.05) | 0.495 |
| IL-2, pg/ml | 2.77(2.32,3.55) | 2.89(2.45,3.54) | 2.97(2.45,3.59) | 0.731 |
| IL-4, pg/ml | 2.17(1.87,2.61) | 2.45(2.05,2.70) | 2.14(1.89,2.46) | 0.213 |
| IL-6, pg/ml | 18.4(10.4,53.0) | 18.9(13.7,60.1) | 23.7(15.3,56.4) | 0.941 |
| IL-10, pg/ml | 6.7(3.9,8.1) | 6.9(5.8,9.0) | 6.7(3.9,8.1) | 0.123 |
| FER, µg/L | 787(300,1057) | 528(300,1253) | 793(306,985) | 0.976 |

Data are n (%), or median (interquartile range). 2019-nCoV, 2019 novel coronavirus; SBP, Systolic Pressure; DBP, diastolic pressure; SpO<sub>2</sub>, oxygen saturation; WBC, white blood cell PMN, neutrophil; LYM, lymphocyte; CRP, C-reactive protein, PCT, procalcitonin; IL-2, interleukin-2; IL-4, interleukin-4; IL-6, interleukin-6; IL-10, interleukin-10; FER, ferroprotein

Oxygen therapy, antiviral therapy(a-interferon, Kaletra[lopinavir/ritonavir]), immunoenhancement therapy (thymosin), prevention of bacterial infection, relieving cough eliminating phlegm and nutritional support were commonly used for all of the 46 patients; while, 26 of them received extra low-dose methylprednisolone treatment with the dosage of 1-2mg/kg/d for 5-7 days via intravenous injection. The specific dosage and duration of methylprednisolone for the patients were determined according to the clinical manifestations, leucocyte count, lymphocyte count, inflammatory index and lesion range. There was no significant difference of age, sex, comorbidities, clinical or laboratory parameters between patients with and without methylprednisolone administration(Table 1). Among the 3 deaths, 2 happened in the patients with methylprednisolone administration and the other one did not receive methylprednisolone treatment.

Among the 46 patients, 27 patients suffered from fever(≥37.3°C) on admission, and 15 patients received methylprednisolone therapy, while the remaining patients not. The dynamic change of the body temperature for the 27 patients was shown in Figure 1A. The number and percentage of the patients whose body temperature decreased to the normal range on different day from treatment beginning was reported as follows(methylprednisolone therapy *vs*. no methylprednisolone therapy): day 2, 7(46.7%) *vs*.2(16.7%); day 3, 13(86.7%) *vs*.4(33.4%); day 4, 13(86.7%) *vs*.5(41.7%); day 5, 14(93.3%) *vs*.7(58.3%); day 6, 15(100%) *vs*.9(75%); day 7, 15(100%) *vs*.7(58.3%); day 8, 15(100%) *vs*.10(83.3%); day 9, 15(100%) *vs*.12(100%); The average number of days required for no fever was significant shorter in the patients with methylprednisolone therapy (2.06±0.28 *vs*. 4.39±0.70, P=0.010, Figure 1B).

**Figure 1.**
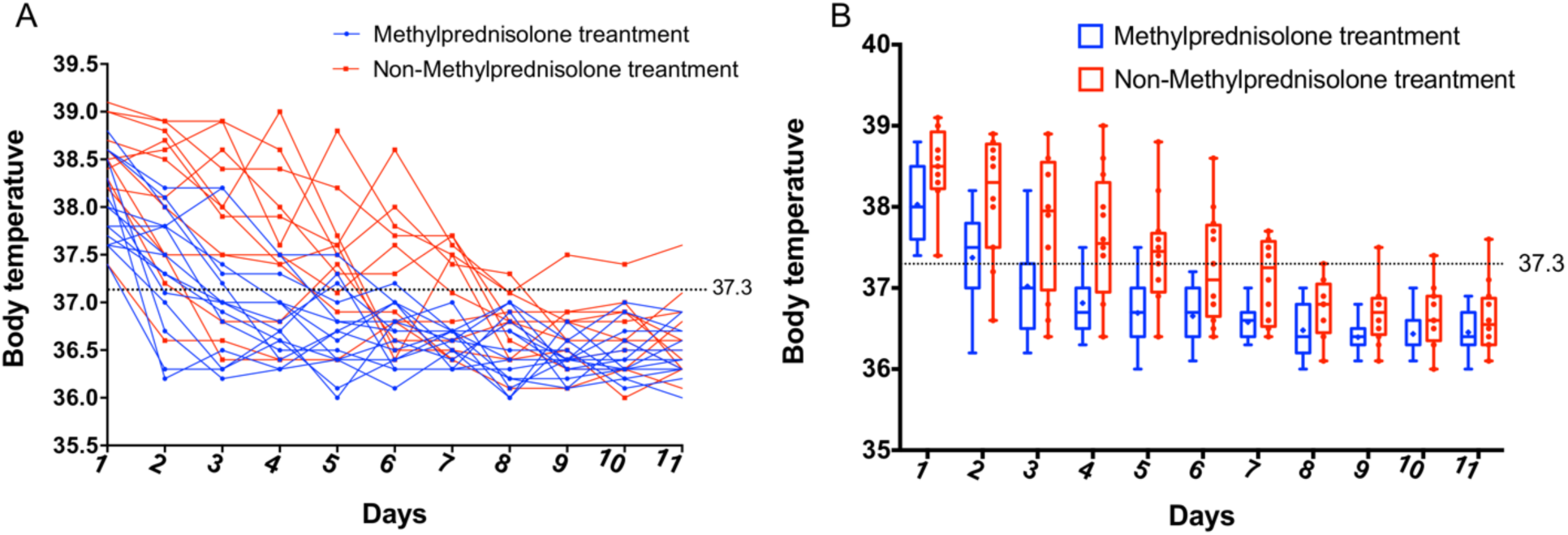
Comparison of the dynamic change of the body temperature between severe COVID-19 pneumonia patients with and without methylprednisolone treatment. (A) the dynamic change of individual patient; (B) comparison for the average and median number.

The median of SpO_2_ at rest was similar between patients with and without methylprednisolone therapy on admission. Due to the low SpO2, all of the 46 patients received oxygen therapy. The daily SpO_2_ of each patient was shown in Figure 2A. The average SpO_2_of every day was compared between patients with and without methylprednisolone therapy, as shown in Figure 3B, the methylprednisolone therapy group had a faster improvement of SpO_2_: day 2, 11(42.3%) *vs*.4(20.0%); day 3, 17(65.4%) *vs*.4(20.0%); day 4, 16(61.5%) *vs*.4(20.0%); day 5, 18(69.2%) *vs*.7(35.0%); day 6, 22(84.6%) *vs*.12(60%); day 7, 25(96.2%) *vs*.14(70.0%); Moreover, patients without methylprednisolone therapy had significantly longer interval of using supplemental oxygen therapy than those with methylprednisolone therapy (8.2days[IQR 7.0-10.3] *vs*. 13.5days(IQR 10.3-16); P<0.001).

**Figure 2.**
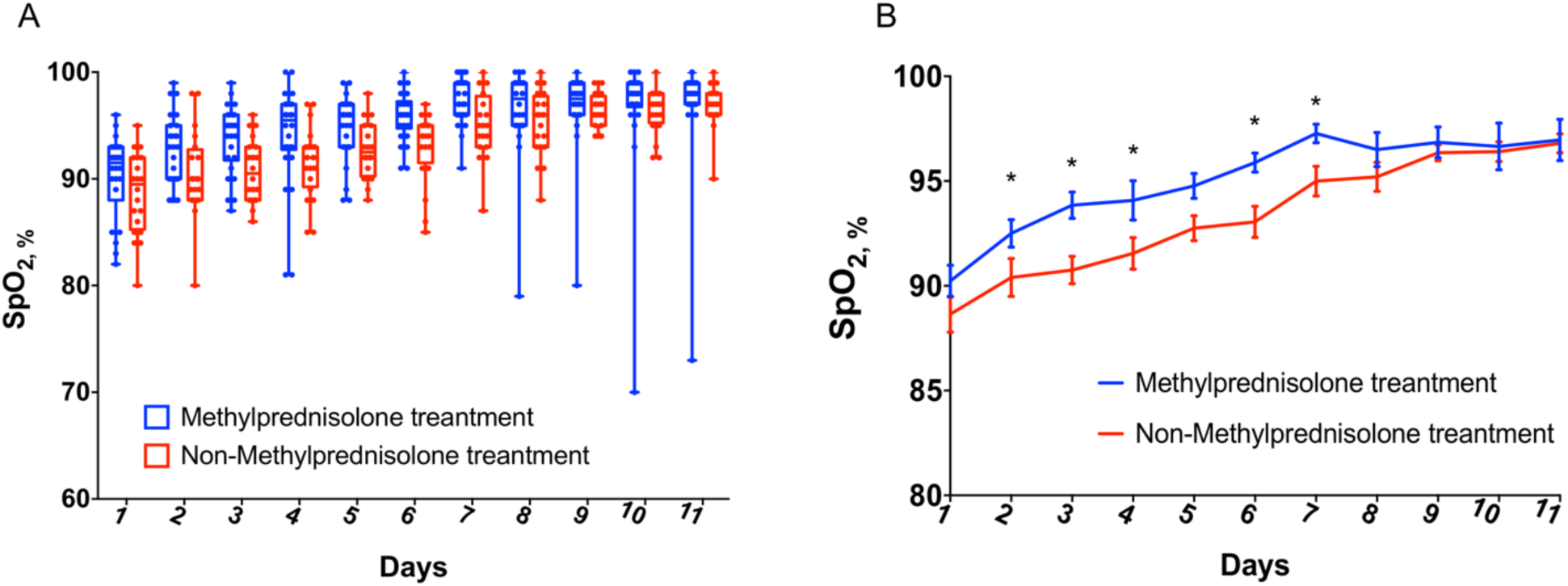
Comparison of the dynamic change of SpO_2_ at rest between severe COVID-19 pneumonia patients with and without methylprednisolone treatment. (A) the dynamic change of individual patient; (B)the trend of SpO_2_ change and comparison for SpO2 on daily basis.

**Figure 3.**
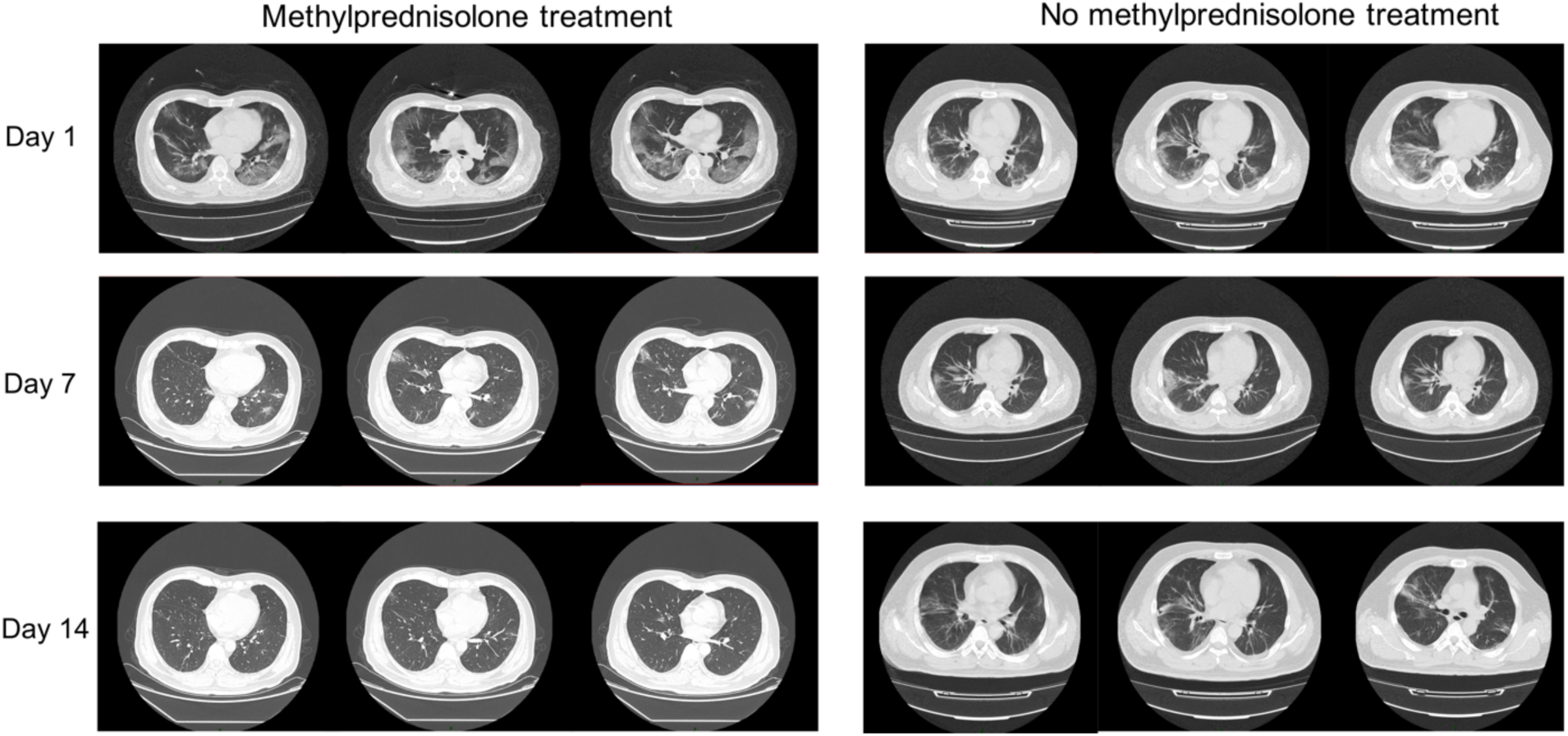
Images of chest CT scan on day 1, 7 and 14 after hospitalization in severe COVID-19 pneumonia patients with and without methylprednisolone treatment

The main laboratory parameters had similar trends between patients with and without methylprednisolone therapy during the hospitalization. The WBC, PMN, LYM, CPR, PCT 0.55, IL-2,IL-4, IL-6 and IL-10 were not significantly different between the two groups on day 6 after the treatment (Table 2). However, in terms of chest CT scan on day 7 and 14, we found the absorption degree of the focus was significantly better in the patients with methylprednisolone (Figure 4).

**Table 2.** Comparison of the main laboratory parameters between severe COVID-19 pneumonia patients with and without methylprednisolone treatment on the sixth day following treatment

|  | Methylprednisolone treatment |  |  |
| --- | --- | --- | --- |
|  | Yes(n=26) | No(n=20) | P value |
| WBC, G/L | 5.22(3.71,8.31) | 6.83(4.21,8.78) | 0.556 |
| PMN,G/L | 3.67(2.82,6.11) | 4.71(3.02,5.87) | 0.335 |
| LYM,G/L | 1.31(0.98,1.91) | 1.43(1.03,1.97) | 0.119 |
| CPR, mg/L | 22.1(9.8,32.3) | 17.4(10.7,56.3) | 0.332 |
| PCT, mg/L | 0.20(0.13,0.79) | 0.19(0.13,0.66) | 0.899 |
| IL-2, pg/ml | 2.53(2.25,2.84) | 2.33(2.04,3.19) | 0.318 |
| IL-4, pg/ml | 1.59(1.38,1.97) | 1.58(1.39,1.81) | 0.634 |
| IL-6, pg/ml | 3.53(3.07,5.05) | 4.11(2.89,7.13) | 0.124 |
| IL-10, pg/ml | 4.15(2.81,6.78) | 5.33(2.43,8.1) | 0.653 |

Data are median (interquartile range). 2019-nCoV, 2019 novel coronavirus; WBC, white blood cell PMN, neutrophil; LYM, lymphocyte; CRP, C-reactive protein, PCT, procalcitonin; IL-2, interleukin-2; IL-4, interleukin-4; IL-6, interleukin-6; IL-10, interleukin-10; FER, ferroprotein

## Discussion

The pathological process of severe COVID-19 pneumonia is the inflammation reaction characterized by destruction of deep airway and alveolar[6]. It is currently considered that the lung injury is not only associated with direct virus-induced injury, but also COVID-19 invasion triggers the immune responses that lead to the activation of immune cells(monocyte, macrophage, T and B-lymphocyte) to release a large number of pro- and anti-inflammatory cytokines including TNF, IL-1β, IL-6 and so on. Overwhelming secretion of cytokines causes severe lung damage, which manifest as extensive damage of pulmonary vascular endothelial and alveolar epithelial cells as well as increased pulmonary vascular permeability, leading to the pulmonary edema and hyaline membrane formation[11,12]. Histologic examination has shown diffuse alveolar damage and mucinous exudate, which similar to acute respiratory distress syndrome[6].

For the patients of severe COVID-19 pneumonia, aggravation of symptoms always occurs during 5-7 days after onset[13]. Therefore, it is important to strengthen the treatment for suppression of pro-inflammatory response and control of cytokine storm at this stage. A majority of patients can survival and recover if they overcome this period. Corticosteroid are the classical immunosuppressive drugs, which are important to stop or delay the progression of the pneumonia and have been proved to be effective for treatment of ARDS[14,15]. In addition to immunosuppressive activities, corticosteroid have an anti-inflammatory role to reduce systemic inflammatory, decrease exudative fluid in the lung tissue, promote absorption of inflammasome and prevent further diffuse alveolar damage, which can relieve hypoxemia and effectively protect the lung to prevent further progression of respiratory insufficiency[16]. Meanwhile, corticosteroid can induce a decrease in body temperature and help alleviate the poisoning symptom caused by hyperthermia.

In the present study, the patients with severe COVID-19 pneumonia had markedly increased inflammatory markers such as CRP, IL-6 and FER, which signified occurrence of the inflammatory reaction phase. Meanwhile, most patients developed fever, cough, dyspnea and markedly decline in oxygen saturation, which are the early clinical manifestations of ARDS.

Based on these, we believed there was the indictor for corticosteroid admission to treat these patients. Judging from the results, early application of low-dose corticosteroid could improve the treatment effect, presenting as improvement of hypoxia and fever symptom, shorten the disease course, and accelerating focus absorption. Benefited from condition monitoring and refined management, no serious complications caused by corticosteroid happened in these patients.

In addition to the timing of treatment, it is also important to master the treatment duration and choose appropriate corticosteroid formulations and dosage. The basic principles of corticosteroid formulations selection lie in the following two aspects: a short half-life and strong penetrating ability. The corticosteroid formulation used in our cohort was methylprednisolone, a median effect corticosteroid with a half-life of 12-36, which has been proved to be associated with a better intensity of immunosuppression[17]. In our experience, the dosage, duration and route of methylprednisolone administration was 1-2mg/kg/d for 5-7 days via intravenous injection. Nevertheless, the specific dosage and duration for individual patient was determined on the clinical manifestations, leucocyte and lymphocyte count, inflammatory index and lesion range.

Corticosteroid treatment-induced complications is the other main concern. The most common complication caused by corticosteroid is secondary infection(fungi and bacteria)[18]. In our experience, once the secondary infection occurs in patients with severe COVID-19 pneumonia, sensitive and full-dose antibacterial drugs should be immediately added. The majority of patients are prone to secondary bacterial infection due to exudate accumulation in the lungs, that push the prophylactic use of antibiotics such as moxifloxacin, levofloxacin and cephalosporins. Secondly, the use of immune regulators(human immunoglobulin) can enhance the immune function of the patients. In our experience, human immunoglobulin was usually used in the critical patients with a dosage of 10-20 g/d for 7-10 days as the pulse therapy. Thirdly, thymosin secreted by thymic epithelial cells can promote the maturation of T-lymphocytes and regulate function of cellular immunity. We suggest using thymosin during hospitalization and the course of treatment can be determined according to the results of lymphocyte test. Fourthly, ensure adequate caloric, protein and vitamins intake and maintain the electrolyte and acid–base balance. The other common complication is hyperglycemia[19]; therefore, continual blood glucose monitoring is recommended, and antidiabetic agents should be used when necessary. The uncommon complications caused by corticosteroid are gastrointestinal bleeding, hypertension, venous thrombosis, hypokalemia and so on, all of these should be paid attention by us[20]. The prudent use of corticosteroid and preventive measure are essential to prevent deterioration in elder patients with these comorbidities.

This study has several limitations. First, the study was a retrospective single center observational research with a small size and no external validation cohort, therefore, unmeasured confounders may influence the accuracy of the results. Secondly, the study has not included the mid- and long-term outcome of the cohort after discharge, and continued follow-up observation was needed.

In conclusion, our data indicate that in patients with severe COVID-19 pneumonia, early, low-dose and short-term application of corticosteroid was associated with a faster improvement of clinical symptoms and absorption of lung focus.

## Data Availability

The data used to support the findings of this study are included in the article.

## Acknowledgements

This study was financially supported by the Natural Science Foundation of China (NO.81700317).

## Conflict of interest

The authors declare no conflicts of interest.

## Notes

### Competing Interest Statement

The authors have declared no competing interest.

### Funding Statement

This study was financially supported by the Natural Science Foundation of China (NO.81700317 to Y.W).

